# Inequities in COVID-19 vaccine and booster coverage across Massachusetts ZIP codes: Large gaps persist after the 2021/22 Omicron wave

**DOI:** 10.1101/2022.04.07.22273593

**Authors:** Jacob Bor, Sabrina A. Assoumou, Kevin Lane, Yareliz Diaz, Bisola Ojikutu, Julia Raifman, Jonathan I. Levy

## Abstract

**Background:** Inequities in COVID-19 vaccine coverage may contribute to future disparities in morbidity and mortality between Massachusetts (MA) communities.

**Methods:** We obtained public-use data on residents vaccinated and boosted by ZIP code (and by age group: 5-19, 20-39, 40-64, 65+) from MA Department of Public Health. We constructed population denominators for postal ZIP codes by aggregating Census-tract population estimates from the 2015-2019 American Community Survey. We excluded non-residential ZIP codes and the smallest ZIP codes containing 1% of the state’s population. We mapped variation in ZIP-code level primary series vaccine and booster coverage and used regression models to evaluate the association of these measures with ZIP-code-level socioeconomic and demographic characteristics. Because age is strongly associated with COVID-19 severity and vaccine access/uptake, we assessed whether observed socioeconomic and racial inequities persisted after adjusting for age composition and plotted age-specific vaccine and booster coverage by deciles of ZIP-code characteristics.

**Results:** We analyzed data on 418 ZIP codes. We observed wide geographic variation in primary series vaccination and booster rates, with marked inequities by ZIP-code-level education, median household income, essential worker share, and racial-ethnic composition. In age-stratified analyses, primary series vaccine coverage was very high among the elderly. However, we found large inequities in vaccination rates among younger adults and children, and very large inequities in booster rates for all age groups. In multivariable regression models, each 10 percentage point increase in “percent college educated” was associated with a 5.0 percentage point increase in primary series vaccine coverage and a 4.9 percentage point increase in booster coverage. Although ZIP codes with higher “percent Black/Latino/Indigenous” and higher “percent essential workers” had lower vaccine coverage, these associations became strongly positive after adjusting for age and education, consistent with high demand for vaccines among Black/Latino/Indigenous and essential worker populations.

**Conclusion:** One year into MA’s vaccine rollout, large disparities in COVID-19 primary series vaccine and booster coverage persist across MA ZIP codes.

**Key Messages**

- As of March 2022, in the wake of MA’s Omicron wave, there were large inequities in ZIP-code-level vaccine and booster coverage by income, education, percent Black/Latino/Indigenous, and percent essential workers.
- Education was the strongest predictor of ZIP-code vaccine coverage in MA.
- Coverage gaps in ZIP codes with many essential workers and large Black/Latino/Indigenous populations are troubling, as these groups face disproportionate risk for COVID-19 infection and severe illness. However, we found no evidence that “hesitancy” drives vaccination gaps. After adjusting for age and education levels, vaccine uptake was higher in ZIP codes with many Black/Latino/Indigenous residents or essential workers.
- Gaps in vaccine and booster coverage among vulnerable groups may lead to excess morbidity, mortality, and economic losses during the next COVID-19 wave. These burdens will not be equitably shared and are preventable.

## BACKGROUND

Vaccination against coronavirus disease 2019 (COVID-19) – including the recommended booster shot – is a critical line of defense against severe illness, hospitalization, and death. Communities with low vaccination rates may be particularly vulnerable to future waves of the COVID-19 epidemic^1,2^. While vaccine uptake in Massachusetts (MA) is high relative to the national average, there remain an appreciable number of eligible individuals who remain unvaccinated, and an even greater number who have not received boosters. A crucial question is whether there are inequities in the distribution of vaccination or booster rates that could point toward the need for targeted public policy measures.

Nationally, vaccination rates are lower for less educated and more rural communities and vaccine uptake has been slower in Black and Hispanic populations.^3,4^ Although “vaccine hesitancy” dominates media coverage, in fact, language barriers, lack of regular health providers, absence of paid time off to get vaccinated and recover, and lack of trust in the health system all play a role in undermining vaccine coverage^5,6^.

In February 2021, the Baker-Polito Administration launched the MA Vaccine Equity Initiative (VEI) to address vaccine hesitancy and improve vaccine administration rates in the twenty most disproportionately impacted communities.^7^ These communities were identified based on COVID-19 case rates, CDC-defined social vulnerability, and the share Black, Indigenous, and people of color.^8^ These populations, as well as the overlapping population of essential workers, have been at elevated risk for COVID exposure, infection, morbidity, and mortality throughout the pandemic^9–15^. The VEI allocated has awarded $46.5 million to 167 community organizations to support vaccine education and outreach.^16^

In this report, we assess variation in primary vaccine series and booster coverage across Massachusetts ZIP codes, analyzing data after the Winter 2022 Omicron variant wave and one year into MA’s population vaccine rollout. Boosters are a critical line of defense against the Omicron variant and will likely be important in mitigating impacts of future waves^17,18^. ZIP-codes are small enough to capture socio-demographic heterogeneity that is obscured in city, town, or county-level estimates. ZIP-code data also enable identification of areas with low vaccine uptake where the consequences of future COVID-19 waves could be most dire. We additionally stratify our analysis by age, the leading risk factor for severe illness due to COVID-19^19^ and a strong correlate of vaccination rates. Our analysis complements aggregate reporting by the MA Department of Public Health (DPH) on the 20 cities and towns that have been the focus of the VEI, as well as prior reports of vaccination patterns at the city/town level and across Boston ZIP codes^20^.

## METHODS

### Data Sources

#### Vaccines and booster shots delivered

We extracted data on counts of vaccinations by ZIP code and sex and by ZIP code and age group, published by MA DPH as “Weekly COVID-19 Municipality Vaccination Data”.^21^ Data are reported to the state by health facilities and vaccination sites. Residential postal ZIP codes and patient age are extracted from paperwork filed at the vaccination site. We analyzed data on vaccines administered in MA from the start of the vaccine rollout in December 2020 through March 1, 2022.

We extracted data on two key constructs reported in the MA DPH vaccine database:

- Primary vaccine series: one shot if Ad26.COV2.S (e.g. Janssen/Johnson and Johnson), two shots if mRNA-1273 (e.g. Moderna) or BNT162b2 (e.g. Pfizer-BioBTech) vaccine; this definition corresponds with the “fully vaccinated” data reported by MA DPH.
- Booster shot (any booster shot after completing initial vaccine schedule)

At the start of 2022, all MA residents ages 5 and over were eligible to be vaccinated, and all residents ages 12 and over were eligible for the booster. Individuals were eligible for the booster 6 months after the second dose of mRNA-1273 or BNT162b2 – reduced to 5 months on January 4 – or 2 months after their single-dose of Ad26.COV2.S.^22^

MA DPH reports data on vaccines administered by ZIP-code, stratified by age, sex, and race. ZIP-code-level totals, which are not reported, can be constructed through aggregation. To protect confidentiality, MA DPH suppresses exact numbers in cells with fewer than 30 people vaccinated/boosted. To minimize the influence of missing data in the ZIP-code-by-age data, we instead used ZIP-code-by-sex data to estimate aggregate ZIP-code totals for people ages 5 and older. The ZIP-code-by-sex data were nearly complete for self-identified “males” and “females”. Data for the category “neither male nor female” were frequently suppressed due to small numbers (70% of ZIP codes were missing data on this group for vaccines, and 92% were missing data on boosters). We imputed data for “neither male nor female” as follows. We computed the ratio of people vaccinated/boosted among persons “neither male nor female” relative to the number of people vaccinated/boosted among persons either “male” or “female”. We then used this ratio to estimate the number of people “neither male nor female” vaccinated/boosted based on the total number of “male” and “female” people vaccinated/boosted for that ZIP code. We then aggregated across all sexes to obtain counts of ZIP code residents who had received the primary vaccine series and/or booster shot.

In addition to ZIP-code totals, we assessed ZIP-code-by-age primary-series vaccine and booster coverage. Three percent of MA residents receiving primary series vaccines and 2% of MA residents receiving boosters did not report their ZIP code and were excluded from the analysis.

#### Population Denominators

We constructed ZIP-code-by-age population counts using publicly available data from the American Community Survey (ACS) 5-year combined estimates (2015-2019).^23^ The ACS is a random sample survey of approximately 3 million people in the U.S. population each year conducted by the U.S. Census Bureau. Because postal ZIP codes (reported on vaccine forms) differ from Census-defined ZIP-code tabulation areas (ZCTAs), we constructed population denominators *de novo*, aggregating up from Census tracts. Age-specific population data were extracted from ACS at the Census-tract level. We used the Housing and Urban Development tract to zip code 2019 4^th^ quarter crosswalk to assign these census tract populations to postal ZIP codes.^24^ Most census tracts are fully contained within single ZIP codes; however, some are split across ZIP codes. For these, we allocated age-specific population counts to ZIP codes proportionately based on the populations of the underlying census blocks and their ZIP code membership.

#### ZIP-code Characteristics

Census tract level ACS 5-year estimates for 2019 were obtained race/ethnicity (B03002), median household income (B19001), educational attainment and (B15003). Essential workers were defined based on definitions developed by the American Civil Liberties Union, which identified those job types considered “essential” during the pandemic such as work in healthcare, transportation, and food preparation^25^ and has been used to assess COVID-19 inequities in MA.^13–15^ We aggregated to postal ZIP codes using the tract-to-ZIP crosswalk, weighting by tract population. Using the Census ACS estimates we derived zip code level “percent college graduates over the age of 25 years”, “percent Black, Latino, or Indigenous”, “percent essential workers”, and “median household income”. We aggregated Black, Latino, and Indigenous MA residents into a single measure due to the high concentration of each of these groups in a relatively small number of ZIP codes. Aggregation of income data yielded population-weighted averages of median household income across Census tracts within each ZIP code. For convenience, we refer to this ZIP-code level measure as “median household income”. We additionally constructed a series of variables to capture differences in age composition across ZIP codes: % of ZIP-code residents aged 0-4 years, % 5-19 years, % 20-39 years, % 40-64 years, % 65+ years. Finally, we created an indicator for whether the ZIP code was within one of the state’s 20 VEI communities: Boston, Brockton, Chelsea, Everett, Fall River, Fitchburg, Framingham, Haverhill, Holyoke, Lawrence, Leominster, Lowell, Lynn, Malden, Methuen, New Bedford, Randolph, Revere, Springfield, and Worcester.

#### Exclusions

We excluded ZIP codes that did not correspond to residential areas, ZIP codes that corresponded to specific universities or businesses, and ZIP codes assigned to post office boxes and not to residential addresses. This reduced the total number of ZIP codes from 648 to 481. To avoid unstable estimates due to very small denominators, we excluded the smallest ZIP codes containing 1% of the total population (n=63). For the remaining data (n=418), we conducted complete case analyses, excluding ZIP-by-age cells with missing data.

### Analysis

We constructed estimates of “percent vaccinated” (primary series) and “percent boosted” for MA ZIP codes and for ZIP-code-by-age-group cells. Different age categories were reported in the state vaccination data and the ACS. To harmonize age groups and reduce the number of small cells, we collapsed age to the following categories: 5-19 years, 20-39 years, 40-64 years, 65+ years.

Uncertainty exists when estimating vaccine coverage for specific ZIP codes due to sampling error in the population denominators. The ACS population estimates are published with standard errors, however, these are reported at the census tract (not ZIP code) level. We captured this uncertainty when reporting estimates for specific ZIP-codes (and ZIP-by-age cells) by constructing 90% confidence intervals (CI) using a resampling approach. For each census tract by age group cell, we simulated 101 population estimates from a normal distribution defined by the ACS point estimates and standard error for that observation. We then aggregated to ZIP codes and computed the constructs of interest holding the numerator constant. To capture uncertainty in the estimates, we then ranked these simulated estimates and used the 5^th^ and 95^th^ ranked estimate as the lower- and upper-bound for our 90% CI.

#### ZIP-code level analysis

We assessed spatial heterogeneity in vaccine coverage by mapping the percent of residents ages 5 years and older who have received the primary series vaccine or booster shot. We used the same denominator for primary series vaccine and booster coverage to enable comparisons even though children ages 5-11 years were not yet eligible for the booster shot. We created higher-resolution maps for densely populated areas. We then investigated the association of ZIP code vaccination and booster coverage with ZIP code characteristics: median household income, percent college graduates, percent Black/Latino/Indigenous, percent essential worker. We present scatter plots and estimate bivariate and covariate-adjusted associations in linear regression models with robust standard errors. A key aim of the multiple regression models is to determine whether the bivariate associations are explained by other factors.

#### ZIP-code-by-age analysis

To illustrate heterogeneity in age-specific primary series vaccination and booster rates, we plotted ranked coverage rates, ordered by ZIP-code. In these distribution plots, we censored the top and bottom 5% of observations to facilitate visualization of the rest of the distribution. To assess the association between vaccination rates and ZIP-code population characteristics, we constructed population-weighted deciles for each characteristic. We then computed binned averages and 95% CI’s within each decile to estimate the percent vaccinated and percent boosted. We also assessed the association of these ZIP-code characteristics with percent vaccinated and boosted in age-stratified multivariate regression models, allowing for different relationships within each age group.

## RESULTS

We analyzed data on 418 ZIP codes containing 97% of the MA population (**Table 1**). Of these ZIP codes, 184 (44%) were in cities or towns containing multiple ZIP codes. The mean population of the included ZIP codes was 15,967 (range 2,014 to 61,099) individuals. On average, the study population resided in ZIP codes where 41% of residents were college educated, 19% were Black, Latino, or Indigenous, 32% were essential workers, and where median household income was $41,100.

**Table 1.** Data description.

|  |  |
| --- | --- |
| ZIP codes (N) | 418 |
| Total population (N) | 6,674,243 |
| ZIP code population (mean) | 15,967 |
| ZIP code population (range) | 2,014 to 61,099 |
| *% with a primary series vaccine* | *Mean (SD)* |
| Aged 5+ years | 80% (10%) |
| 5-19 years | 60% (17%) |
| 20-39 years | 77% (14%) |
| 40-64 years | 83% (9%) |
| 65+ years | 97% (10%) |
| *% with a booster shot* | *Mean (SD)* |
| Aged 5+ years | 43% (10%) |
| 20-39 years | 35% (12%) |
| 40-64 years | 50% (11%) |
| 65+ years | 74% (10%) |
| *ZIP-code characteristics* | *Mean (SD)* |
| Median HH income | \$41,100 (\$12,500) |
| % college graduates | 41% (17%) |
| % Black, Latino, or Indigenous | 19% (20%) |
| % essential workers | 32% (6%) |

Note: Mean (standard deviation) of vaccination coverage and ZIP-code characteristics are weighted by ZIP code population ages 5 years and older.

On average, 80% of residents ages 5 years and older had received the primary series vaccine and 43% had received a booster shot. Ninety-seven percent of residents ages 65 years and older had received the primary series vaccine. Coverage for the primary series vaccine was lower in younger age groups, with 83% of persons 40-64 years, 77% of persons 20-39 years, and 60% of persons 5-19 years vaccinated. Booster coverage was 74% among residents 65 years and older, 50% among persons 40-64, 35% of persons 20-39 years.

The maps in **Figure 1** show ZIP-code-level variation in vaccination (top) and booster (bottom) coverage for residents ages 5 years and older. The panels show similar geographic patterns of vaccine and booster uptake, with the highest coverage among ZIP codes in Boston’s Western suburbs, the North and South Shore, Cape Cod and the Islands, and in the college towns of the Pioneer Valley. Lower vaccine and booster coverage was observed in lower-income ZIP codes of the urban centers – greater Boston, Worcester, Springfield, New Bedford, Fall River, Lawrence, and Lowell (**Appendix Figure 1** shows higher resolution maps of these areas). Additionally, lower coverage was observed in non-urban ZIP codes in Central, Western, and Southeast MA, and in MA ZIP codes along the borders with New Hampshire, Rhode Island, and Connecticut.

**Fig 1.**
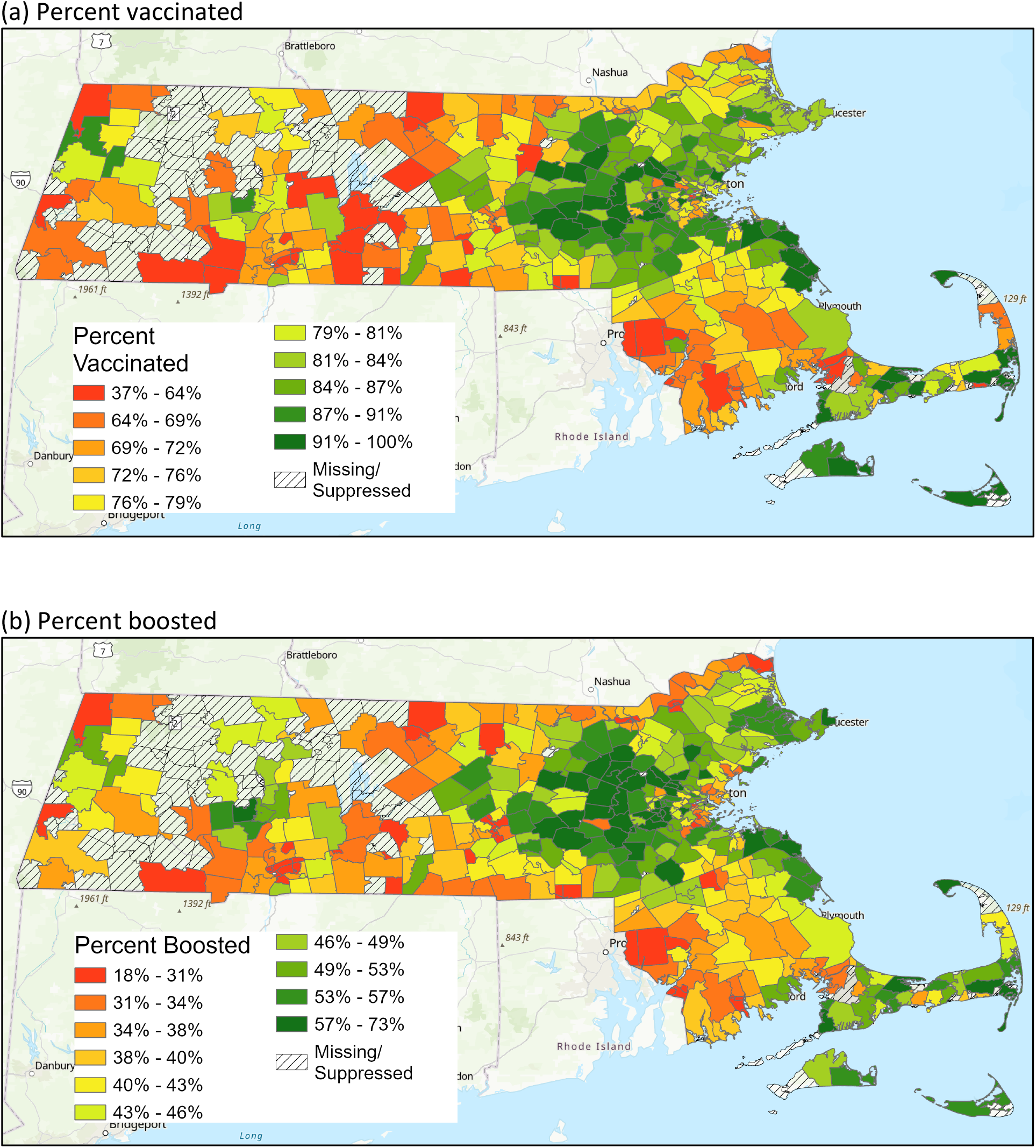
Percentage of residents with a COVID-19 (a) vaccine or (b) booster by ZIP code.

**Figure 2** shows scatter plots of vaccine and booster coverage against ZIP-code characteristics. We observed strong correlations between primary series vaccine and booster coverage and the three sociodemographic indicators. A $10,000 increase in a ZIP code’s median household income was associated with a 4.4 percentage point increase in primary series vaccine coverage and a 6.4 percentage point increase in booster coverage. From the lowest to the highest income levels, booster coverage increased from under 30% to over 60%. A 10 percentage point increase in percent college graduates was associated with a 2.8 percentage point increase in vaccinations and a 4.8 percentage point increase in boosters. And a 10 percentage point increase in percent essential workers was associated with a 5.2 percentage point reduction in primary-series vaccination coverage and a 10.1 percentage point reduction in booster coverage. We observed a weaker relationship between percent Black, Latino, and Indigenous and vaccination coverage, although a 10 percentage point increase in this population share was associated with a 3.0 percentage point decline in booster coverage. Standard errors for these coefficients are shown in **Table 2**.

**Fig 2.**
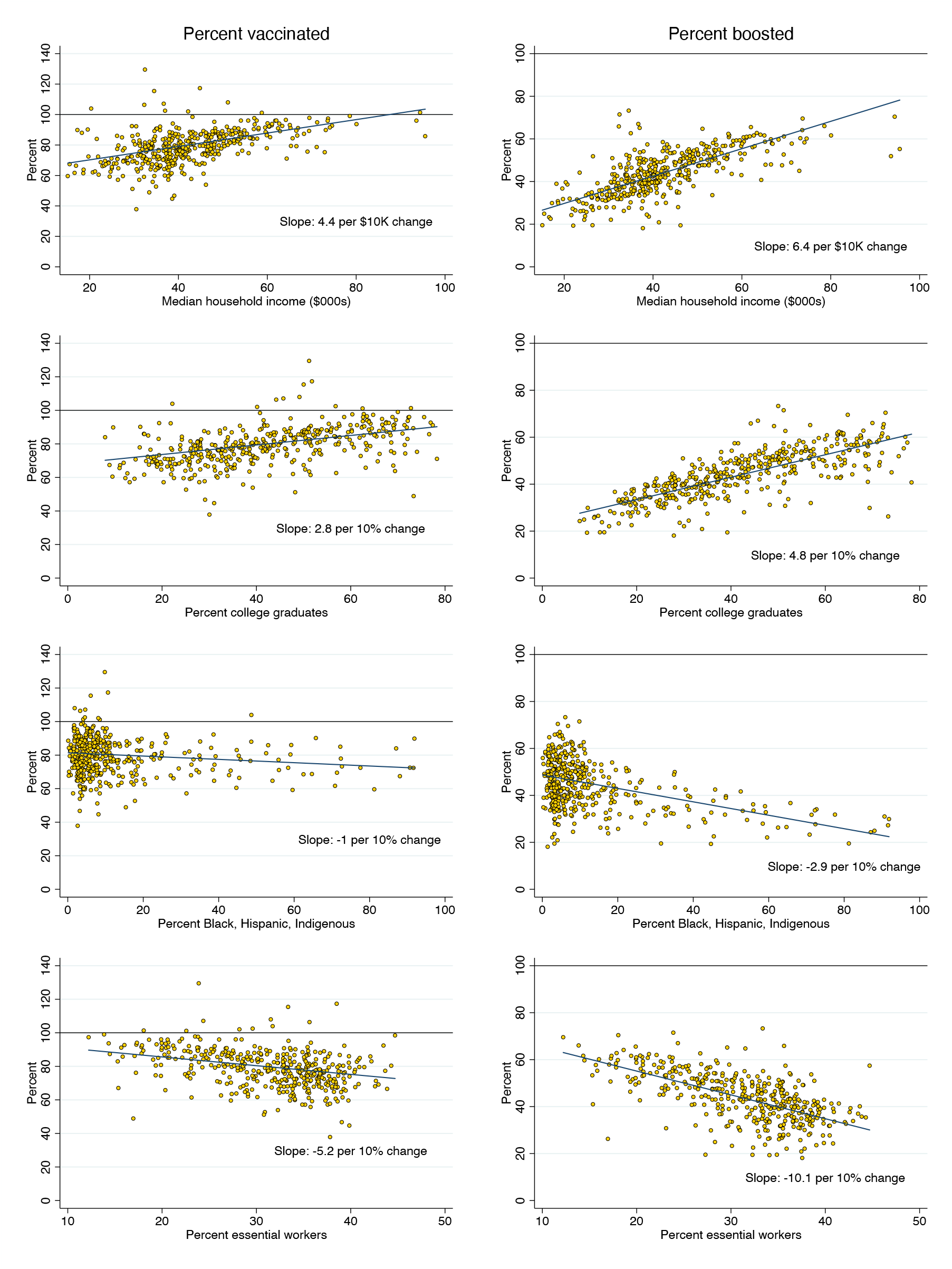
Percent vaccinated and boosted vs. ZIP-code characteristics.

**Table 2.**
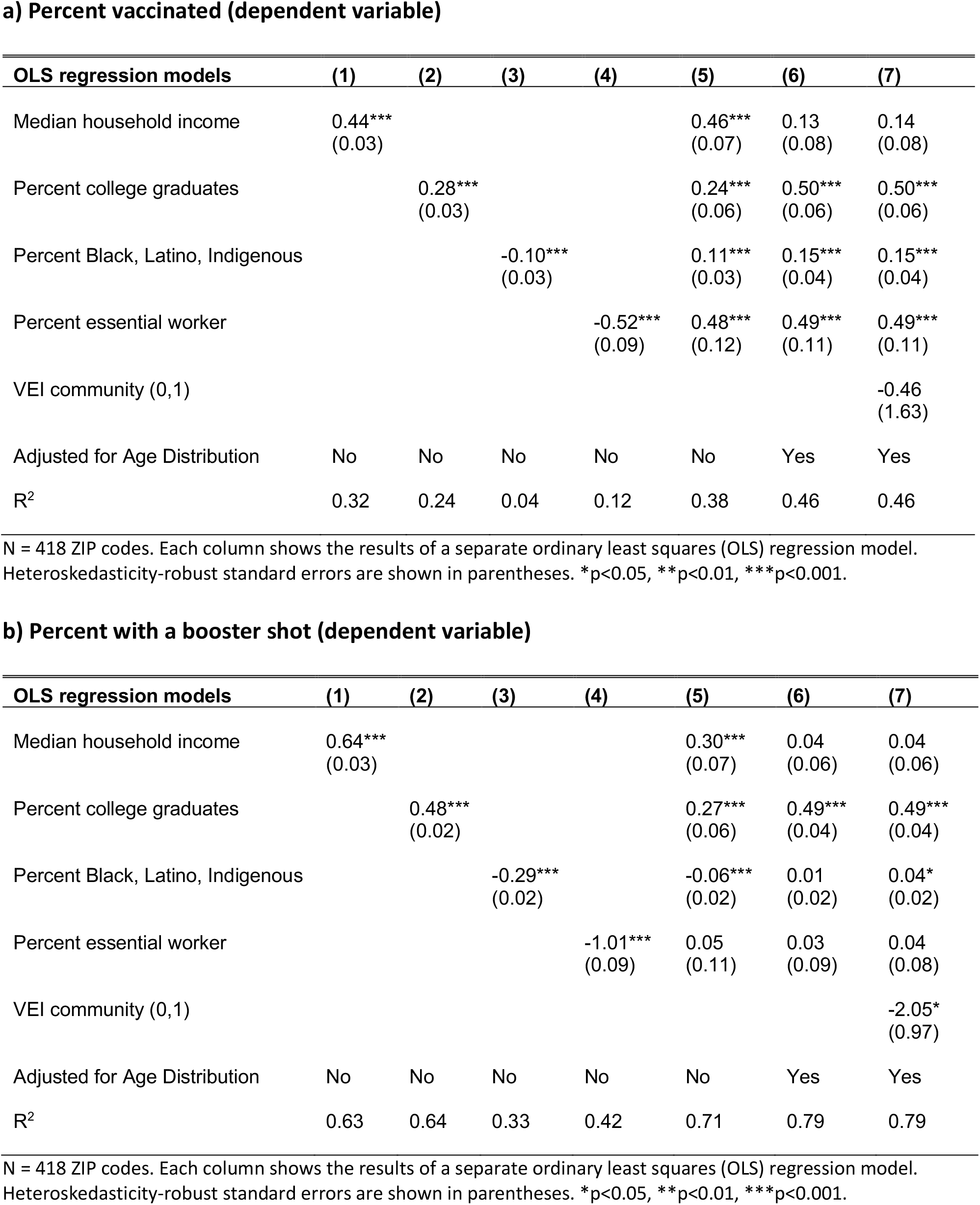
Association of ZIP-code characteristics with the percent of residents aged 5 years and older that are vaccinated COVID-19 and/or have had a COVID-19 booster shot.

Bivariate associations indicate what types of ZIP codes have the highest (or lowest) vaccine coverage. However, because these predictors are correlated, the bivariate associations do not indicate which factors are driving the observed relationships. For example, people who are older tend to have higher incomes and are more likely to be vaccinated; therefore, age composition may confound the relationship between ZIP-code income and vaccine coverage.

**Table 2** shows results from multivariable models regressing percent vaccinated/boosted on variables describing the age composition of the ZIP code (defined above) as well as the four sociodemographic characteristics. In these multivariable regression models, percent college educated emerged as the strongest predictor of vaccine and booster coverage. Each 10 percentage point increase in percent college educated was associated with a 5.0 percentage point increase in vaccine coverage and a 4.9 percentage point increase in booster coverage. Although median household income was strongly positively associated with vaccination (booster) rates in the bivariate setting; this association was attenuated (disappeared) after adjusting for age composition and education.

We observed stark changes in the coefficients on “percent Black, Latino, or Indigenous” and “percent essential workers” after adjusting for age composition and education. In bivariate models, these ZIP-code characteristics were negatively associated with percent vaccinated; however, in multivariable models, the associations turned positive. ZIP codes with a larger share of Black, Latino, and Indigenous residents and with more essential workers had higher vaccination rates than would be otherwise expected based on the education and age composition in those ZIP codes. Adjusting for age and education, each 10 percentage point increase in “percent Black, Latino, or Indigenous” was associated with a 1.5 percentage point increase in vaccine coverage and each 10 percentage point increase in “percent essential workers” was associated with a 4.9 percentage point increase in vaccine coverage. For boosters, while the associations from the bivariate models were similarly attenuated after adjusting for age and education, there was a less pronounced association with percent essential workers (null) and percent Black, Latino, or Indigenous (modestly positive).

The last column of **Table 2** includes an indicator for whether the ZIP code was a part of one of the 20 VEI communities targeted by the state for enhanced vaccine outreach. After adjusting for sociodemographic characteristics and age composition, we find no evidence that VEI communities had higher primary series vaccination than non-VEI communities. VEI communities had slightly lower (−2 percentage points) booster coverage than non-VEI communities after adjusting for sociodemographics. (We note that other factors not included in our model may explain the low vaccination rates in VEI communities, and that vaccination rates might have been even lower in the absence of the VEI.)

We also evaluated whether ZIP-code inequities in vaccination coverage varied by age group. **Figure 3** (left column) shows the distribution of “percent vaccinated” in each ZIP code, stratified by age group. The point estimates contain estimation uncertainty in the denominator, which explains why some percentages exceed 100%. The figure shows very high vaccination rates among the elderly (65+ years), with relatively little variation across ZIP codes. Variation in primary series vaccine coverage was more pronounced for younger age groups, in particular, for children (5-19 years), where vaccination rates vary from as low as 30% to as high as 90%. **Figure 3** (right column) shows “percent boosted” in each ZIP code, stratified by age group. Children are excluded as most are not yet eligible. The share boosted varied widely in all age groups, even for the elderly. For people over 65 years, the share boosted varied from under 50% in the lowest coverage ZIP codes to over 80% in the highest-coverage ZIP codes.

**Fig 3.**
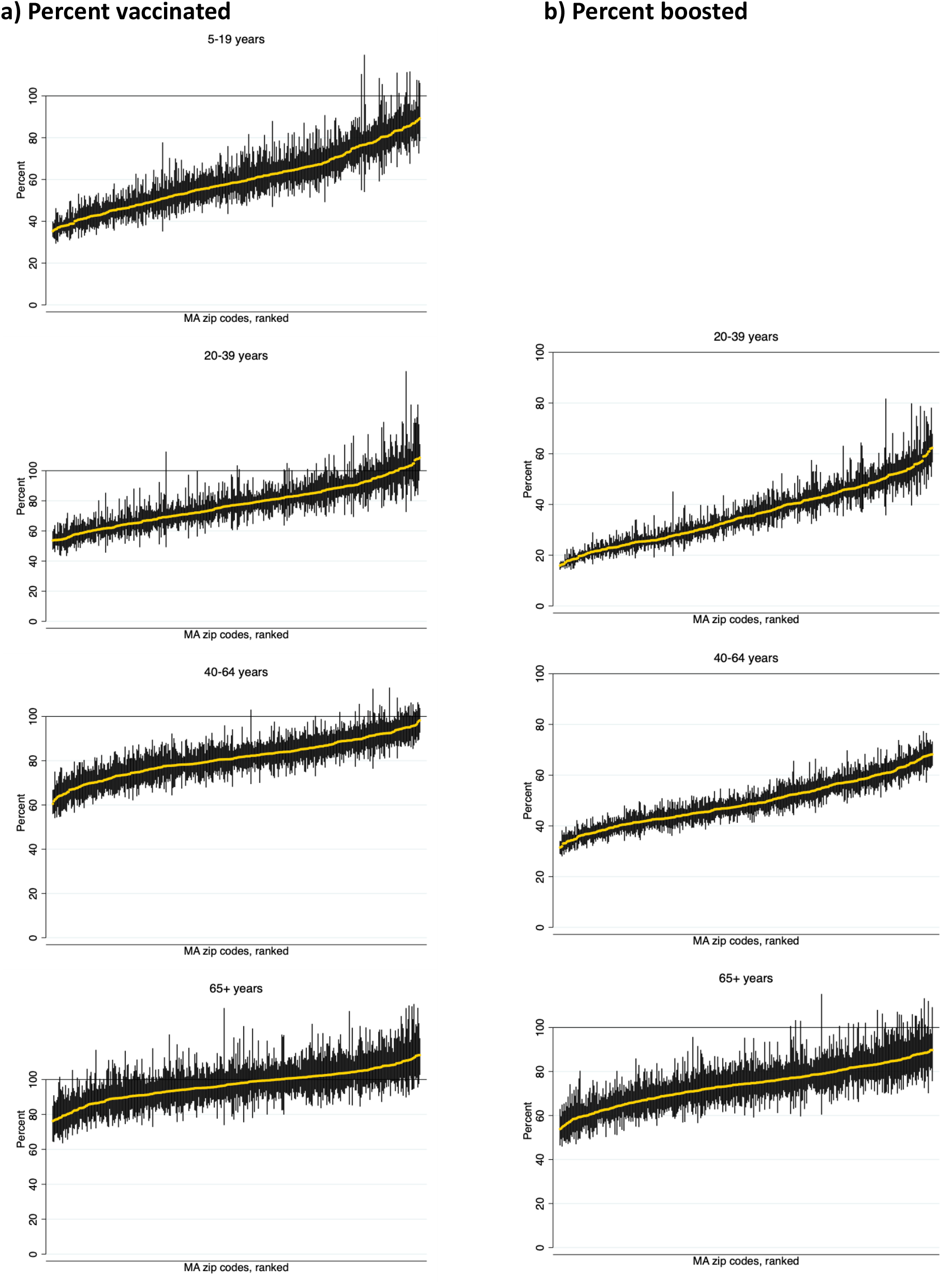
Percentage of residents with COVID vaccine (a) or booster (b), by ZIP code and age. Note: data are ranked by percent vaccinated with the primary series or booster shot.

**Figures 4a-d** show the share of MA adults by age group who are “vaccinated” and “boosted” by the percent of ZIP code residents that are (a) Black, Latino, or Indigenous, (b) college educated, (c) essential workers, and by (d) median household income. MA has achieved very high vaccination coverage among 65+ year-olds across ZIP codes with different population characteristics (bottom-left panel of each figure). Vaccination coverage among MA residents 65+ years is above 90% in all deciles of all four ZIP-code characteristics, and close to 100% for most. The share vaccinated is somewhat lower (90-95%) in the top two deciles of %Black, Latino, or Indigenous and in the bottom four deciles of %college graduates; however, the differences are relatively small. ZIP codes with the largest shares of essential workers had nearly full vaccination coverage among the elderly. Vaccination coverage among adults ages 40-64 years was also relatively high and equitably distributed (middle-left panel of each figure). At least 75% were vaccinated in all ZIP-code covariates deciles, and vaccination rates were *highest* in ZIP codes with a greater share of Black, Latino, or Indigenous residents. Larger gaps (and inequities) in vaccine coverage were apparent for MA adults ages 20-39 (top left panel of each figure).

**Fig 4.**
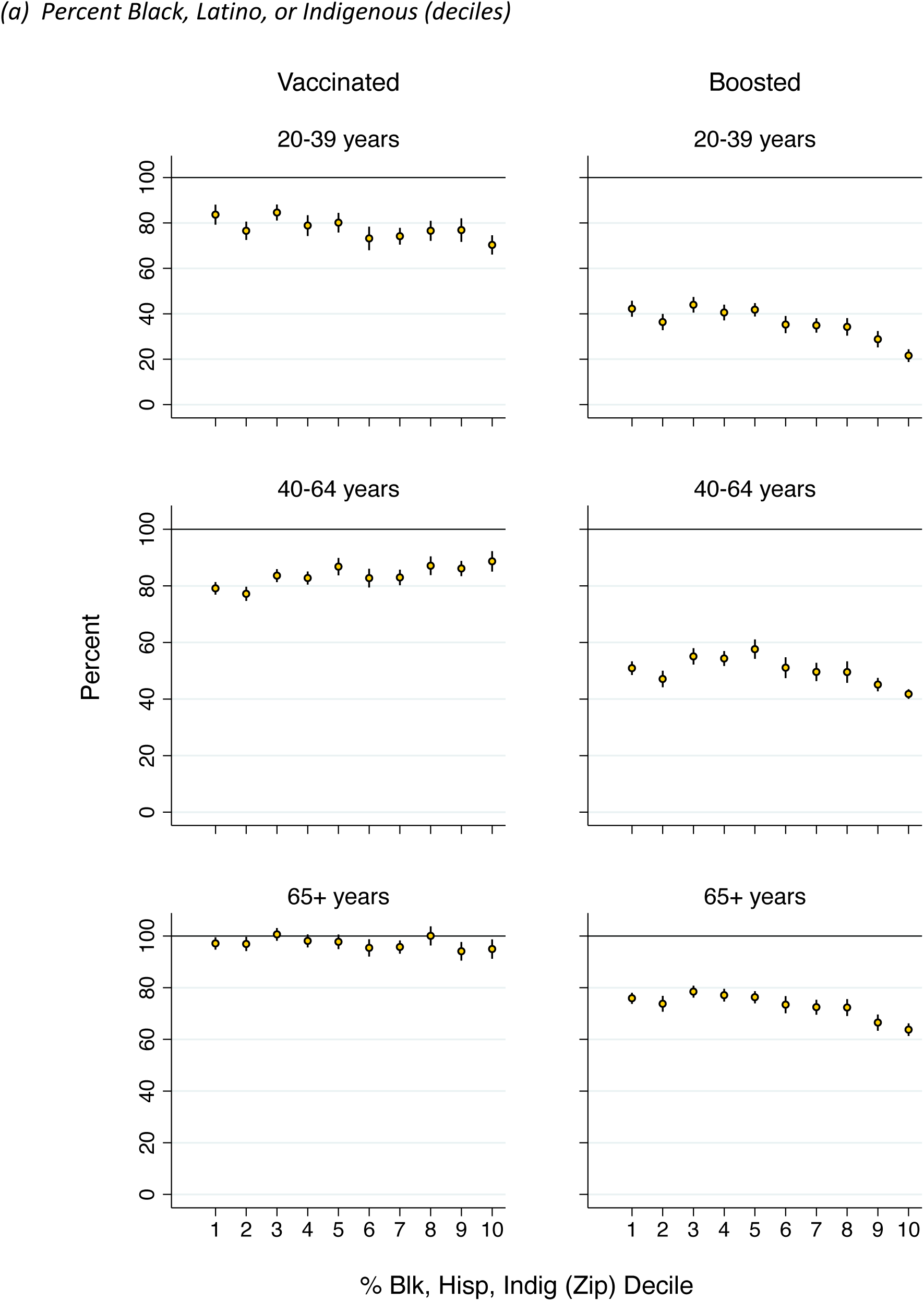
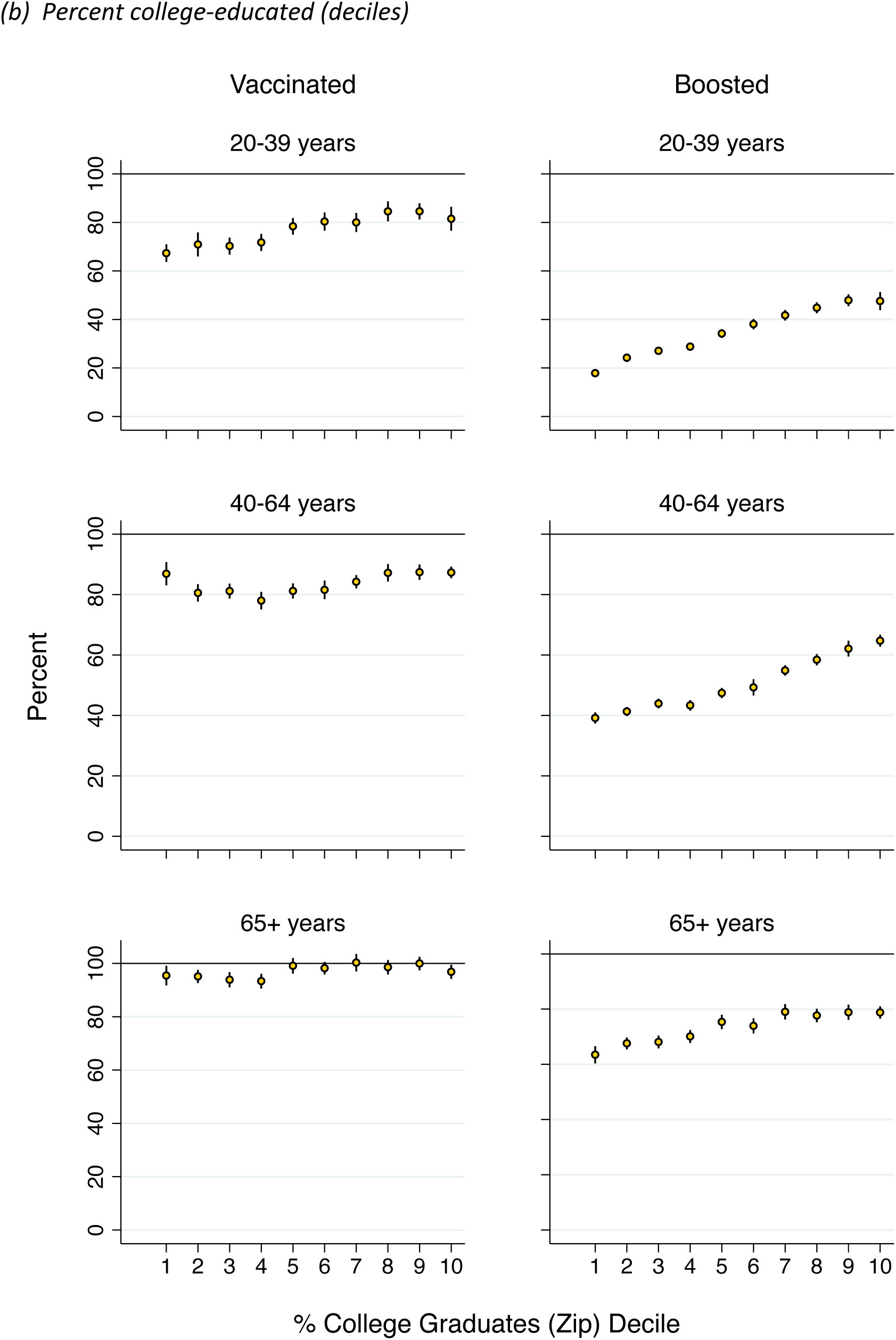
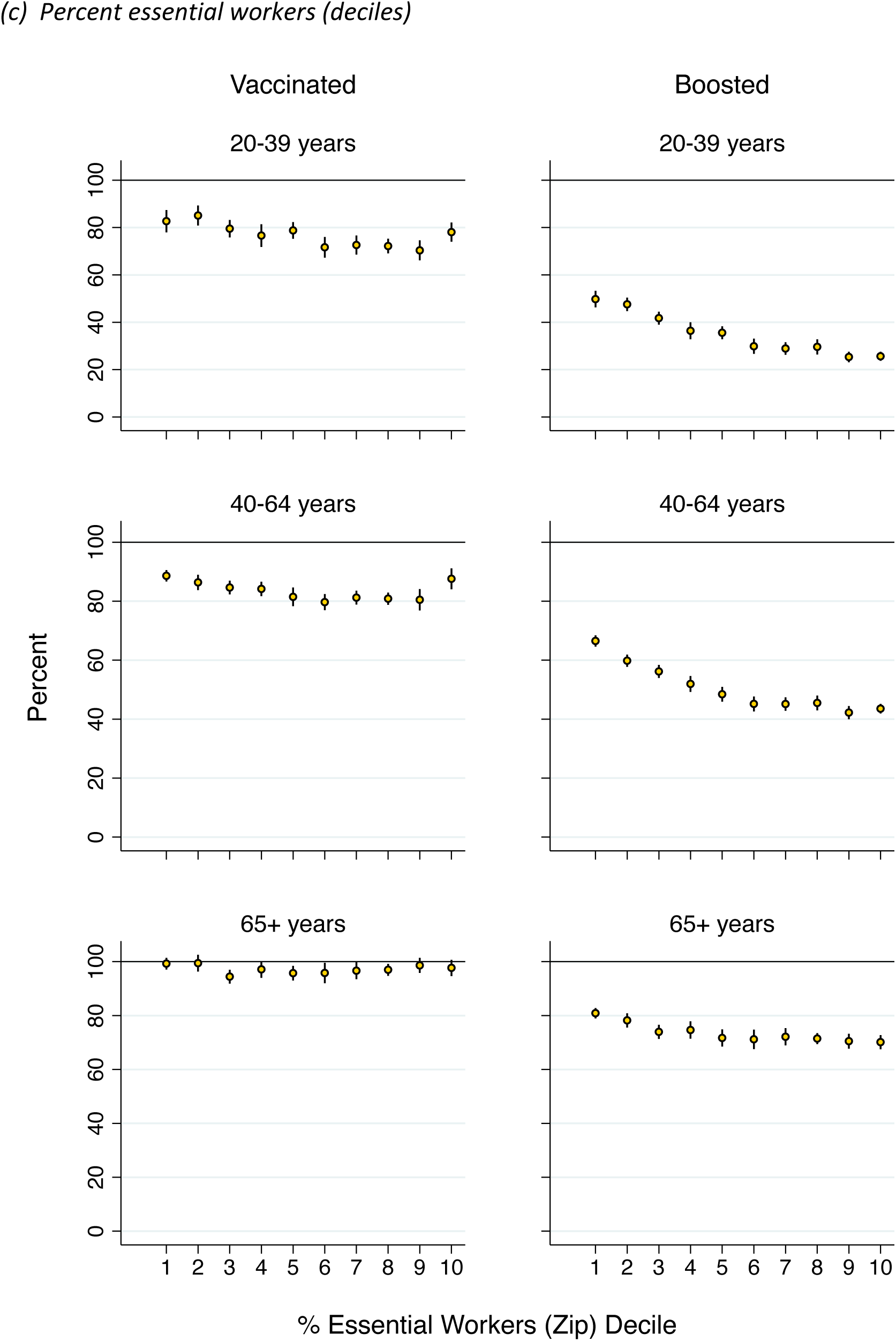
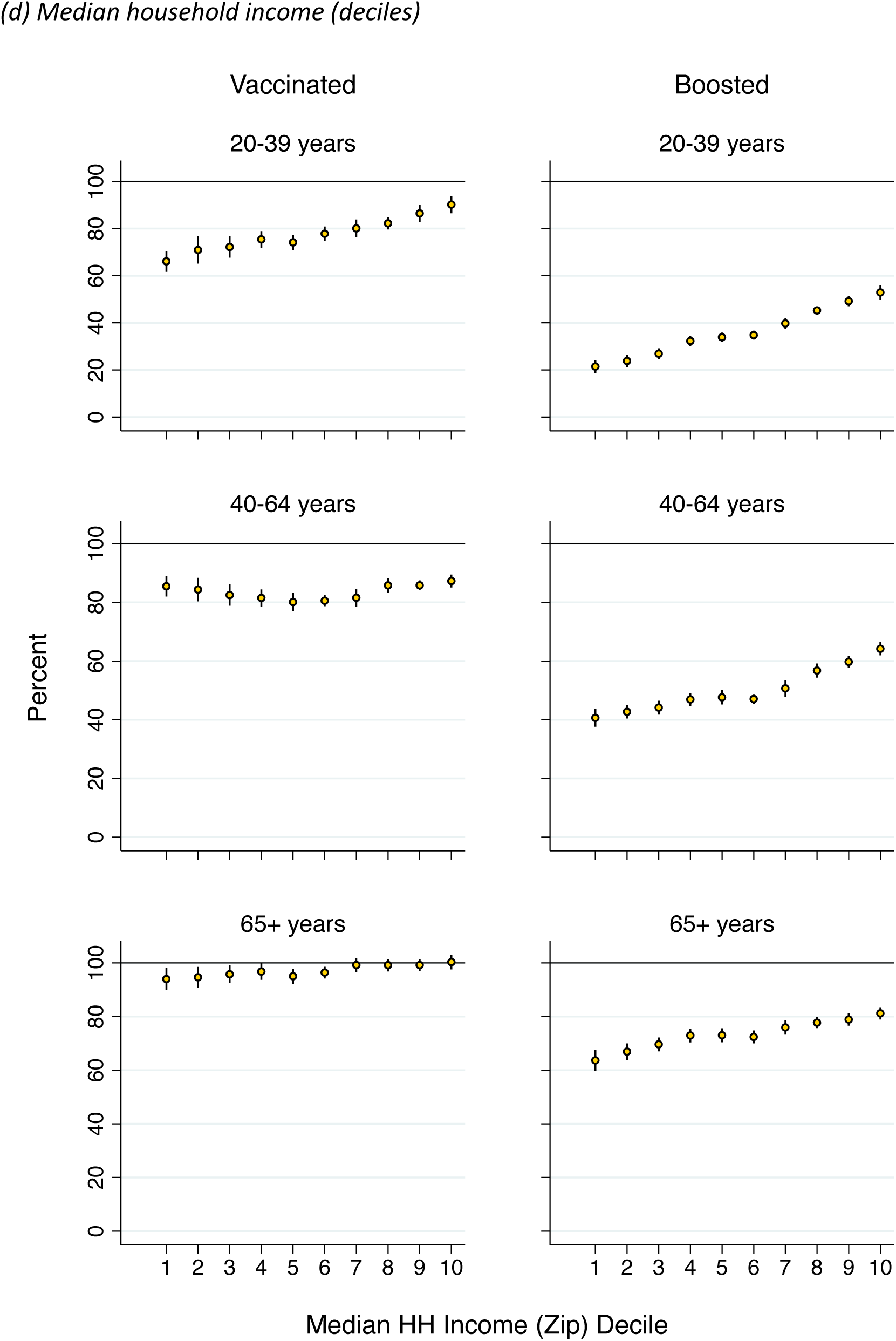
Percent vaccinated (left) or boosted (right) by age and deciles of ZIP-code characteristics.

Despite high, relatively equitable coverage of the original vaccine schedule, there was wide variation in coverage of the vaccine booster (right three panels of each figure). The share of elderly (65+ years) that have received a booster was just over 50% in ZIP codes with the lowest share of college graduates and highest share Black, Latino, or Indigenous; however, booster coverage was over 70% in more-educated ZIP codes and those with larger White populations. Large disparities in booster coverage were also apparent for younger and middle-aged adults, with gaps in coverage of over 25 percentage points between the top and bottom deciles in the ZIP codes stratified by education and by percent essential workers.

**Figure 5** displays the share of MA children (5-19 years) that have been vaccinated. There are large disparities in the share of children who have been vaccinated. About 70% of children have been vaccinated in ZIP codes with the highest share of college educated residents and the lowest share essential workers. Vaccine coverage among children falls with % Black, Latino, and Indigenous, rises with % college educated, and falls with % essential workers. Under 40% of children have been vaccinated in the 10% of ZIP codes with the highest % Black, Latino, and Indigenous and the lowest % college educated.

**Fig 5.**
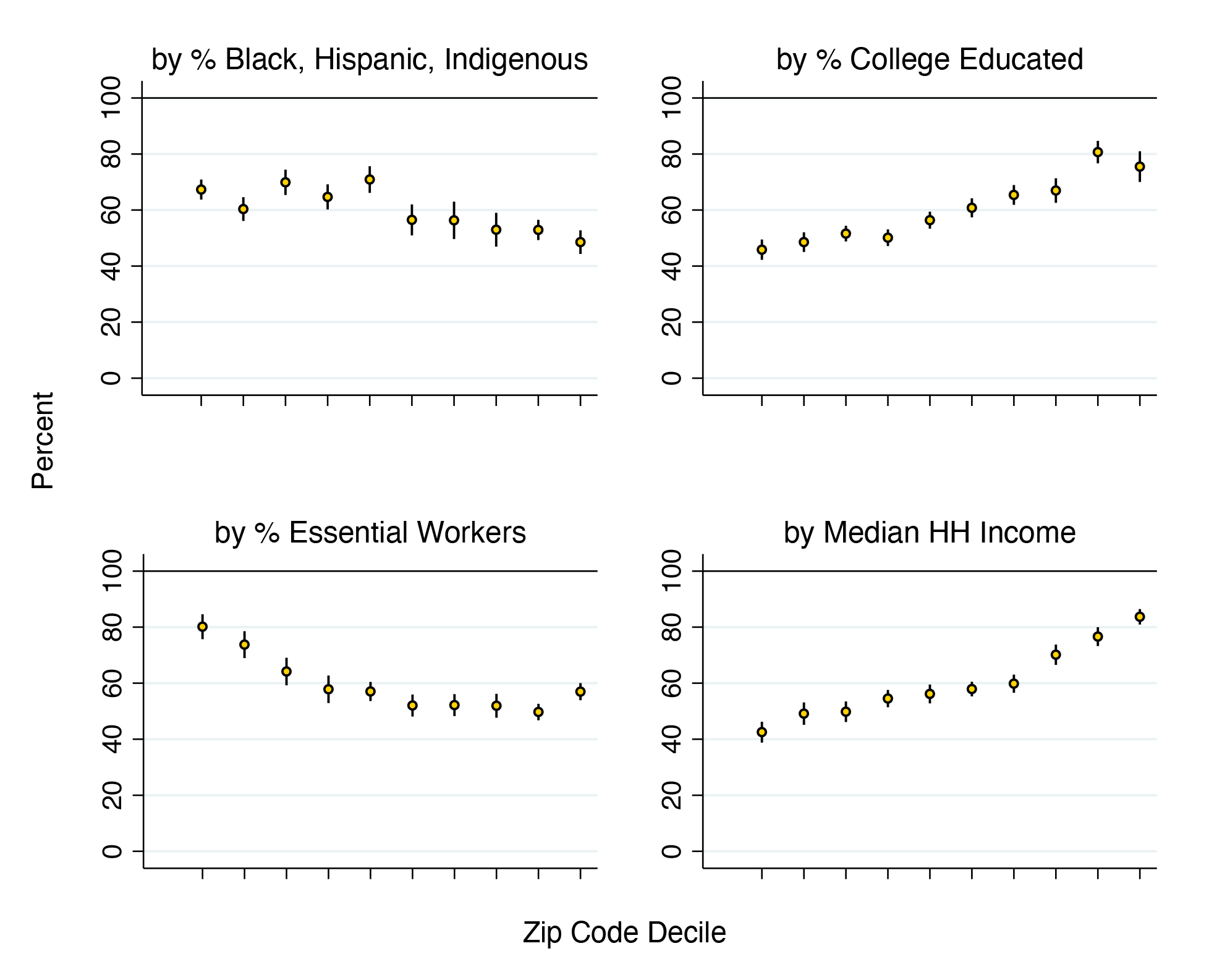
Percentage of MA <u>children vaccinated</u> by ZIP code characteristics (deciles)

**Table 3** presents age-stratified bivariate and multivariable regression models, replicating columns (1)-(4) and (7) of **Table 2** for each age group – 5-19, 20-39, 40-64, 65+ years. In the bivariate models (panel A), income and education had positive associations with vaccine and booster coverage across all age groups. Percent Black, Latino, or Indigenous was negatively associated with vaccine coverage among children and younger adults and with booster coverage at all ages, but positively associated with vaccine coverage among working age adults. Percent essential workers was negatively associated with vaccine coverage for non-elderly adults and children and with booster coverage for all age groups.

**Table 3.** Association of ZIP-code characteristics with percent vaccinated and boosted, by age.

Bivariate regression results*
| *Dependent variable* | **Percent vaccinated** |  |  |  | **Percent boosted** |  |  |
| --- | --- | --- | --- | --- | --- | --- | --- |
| *Age group* | 5-19 | 20-39 | 40-64 | 65+ | 20-39 | 40-64 | 65+ |
| Median household income | 0.97***<br>(0.05) | 0.56***<br>(0.06) | 0.09*<br>(0.04) | 0.14***<br>(0.04) | 0.80***<br>(0.05) | 0.57***<br>(0.03) | 0.37***<br>(0.04) |
| Percent college graduates | 0.63***<br>(0.05) | 0.32***<br>(0.05) | 0.11***<br>(0.03) | 0.09**<br>(0.03) | 0.58***<br>(0.03) | 0.51***<br>(0.02) | 0.29***<br>(0.03) |
| Percent Black, Latino, Indigenous | -0.27***<br>(0.04) | -0.12***<br>(0.04) | 0.13***<br>(0.03) | -0.05<br>(0.03) | -0.30***<br>(0.02) | -0.17***<br>(0.02) | -0.21***<br>(0.02) |
| Percent essential worker | -1.36***<br>(0.14) | -0.52***<br>(0.15) | -0.27***<br>(0.08) | -0.07<br>(0.07) | -1.26***<br>(0.12) | -1.16***<br>(0.06) | -0.51***<br>(0.06) |
| N | 413 | 418 | 418 | 417 | 418 | 418 | 415 |

| *Dependent variable* | **Percent vaccinated** |  |  |  | **Percent boosted** |  |  |
| --- | --- | --- | --- | --- | --- | --- | --- |
| *Age group* | 5-19 | 20-39 | 40-64 | 65+ | 20-39 | 40-64 | 65+ |
| Median household income | 0.95***<br>(0.10) | 0.60***<br>(0.11) | -0.02<br>(0.07) | 0.15<br>(0.09) | 0.41***<br>(0.07) | -0.02<br>(0.06) | 0.11<br>(0.09) |
| Percent college graduates | 0.25**<br>(0.09) | 0.31***<br>(0.07) | 0.37***<br>(0.07) | 0.16*<br>(0.08) | 0.39***<br>(0.04) | 0.55***<br>(0.05) | 0.31***<br>(0.07) |
| Percent Black, Latino, Indigenous | 0.22***<br>(0.05) | 0.13*<br>(0.05) | 0.28***<br>(0.05) | 0.05<br>(0.05) | 0.04<br>(0.03) | 0.12***<br>(0.03) | -0.01<br>(0.04) |
| Percent essential worker | 0.32*<br>(0.15) | 0.74***<br>(0.19) | 0.23<br>(0.14) | 0.47**<br>(0.16) | 0.12<br>(0.12) | -0.08<br>(0.09) | 0.39**<br>(0.13) |
| VEI community (0,1) | -3.85<br>(2.29) | -0.07<br>(2.45) | -0.73<br>(1.87) | -1.64<br>(2.04) | -2.23<br>(1.52) | -2.47<br>(1.28) | -3.79*<br>(1.70) |
| Constant | -2.53<br>(7.54) | 14.37<br>(9.72) | 57.27***<br>(7.11) | 68.91***<br>(8.29) | -1.10<br>(6.36) | 29.97***<br>(4.31) | 45.20***<br>(6.83) |
| R<sup>2</sup> | 0.62 | 0.29 | 0.25 | 0.06 | 0.72 | 0.66 | 0.30 |
| N | 413 | 418 | 418 | 417 | 418 | 418 | 415 |

Each column shows the results of a separate ordinary least squares (OLS) regression model. Heteroskedasticity-robust standard errors are shown in parentheses. \*p<0.05, \*\*p<0.01, \*\*\*p<0.001.

In multivariable models, ZIP code income and education levels maintained strong positive associations with vaccine and booster coverage, with income strongly associated with vaccination of children (5-19) and young adults (20-39 years). After adjusting for income and education, ZIP codes with more essential workers and with larger Black, Latino, or Indigenous populations had similar or higher vaccination coverage across all age groups, in contrast to the bivariate models.

## DISCUSSION

We assessed geographic and sociodemographic equity in the state of Massachusetts (MA)’s coronavirus disease 2019 (COVID-19) vaccination program as of early March 2022, through analysis of newly released data on primary series vaccinations and boosters by age and ZIP code. Our findings indicate that coverage of the primary series vaccine among elderly MA residents (65 years and older) was very high, ranging from 94% in the lowest-income decile of ZIP-codes to 100% in the highest-income decile. This finding likely reflects the state’s consistent emphasis on vaccinating older adults, who are at high risk for severe complications. Primary series vaccine coverage for younger MA residents was lower and exhibited large disparities by ZIP-code-level education, income, percent essential workers, and racial composition. In addition, very large inequities were observed for booster coverage, with gaps of more than 30 percentage points between the lowest- and highest-income ZIP codes. These observed gaps in vaccine-induced protection could have significant adverse health consequences during the next COVID-19 wave, especially given evidence that protection against the Omicron variant is diminished in the absence of a booster.^26,27^

Vaccination coverage gaps are most concerning for populations with high risk for exposure and disease severity if exposed. For example, in the U.S., at the peak of the Omicron BA1 surge, Black people were 3.8 times more likely to be hospitalized than White people.^30^ ZIP codes with larger percentages of essential workers and larger shares of Black, Latino, and Indigenous residents had lower coverage of both the primary series vaccine (for younger adults and children) and the booster (for all age groups). In MA, these groups have experienced disproportionate infection rates, morbidity, and mortality from COVID-19^13–15^ and would therefore be expected to benefit more from vaccination than other groups that are less likely to be exposed to COVID and/or less likely to experience severe illness. State and local public health officials have sought to increase vaccination among Black, Latino, and Indigenous MA residents through the Vaccine Equity Initiative^8^ and to increase vaccination of essential workers through mandates.^28,29^

Despite lower vaccination coverage in communities with large shares of essential workers and Black, Latino, or Indigenous residents, we found that these characteristics were not – after controlling for other ZIP code attributes – associated with lower vaccine uptake. Counter to prevailing assumptions, after adjusting for age composition, income, and education levels, ZIP codes with higher shares of essential workers and Black, Latino, or Indigenous residents had greater uptake of the primary series vaccine. Far from “vaccine hesitancy”^31^, these findings suggest greater demand for vaccination among populations that have been most affected by COVID, after adjusting for age and socioeconomic factors. (Efforts to address medical mistrust may have been important in closing racial disparities in vaccine uptake early in the rollout.) Lagging booster coverage in these groups may reflect timing, as populations vaccinated in Summer and Fall 2021 were not yet eligible for the booster shot at the start of the late 2021 Omicron wave. These findings help reconcile a paradox in the literature which has found lower vaccine coverage among Blacks and Hispanics in aggregate statistics yet greater uptake in individual-level analyses that adjust for covariates.

A causal interpretation of our model would imply that ZIP codes with many essential workers or Black/Latino/Indigenous residents have lower vaccination or booster rates because their populations are on average younger and have lower levels of educational attainment and lower incomes, which may be associated with access. Interventions to address access barriers related to education and poverty – including offering more convenient clinic times, paid sick leave for potential side effects, providing information through trusted community-based sources in multiple languages, and conducting outreach outside of the formal medical sector – will be essential to closing these equity gaps. Local health departments also continue to call for more long-term investments.

These data also show where MA should focus its efforts: vaccinating younger adults 20-39 years, vaccinating children (5-19), and ensuring high, equitable booster coverage among middle-aged and older adults. Recent data indicate that vaccines for children are effective in reducing hospitalizations.^32^ Gaps in booster coverage were very large, with ZIP-code education levels explaining much of the variation. In contrast to the primary vaccine series, ZIP-codes with many essential workers or many Black, Latino, or Indigenous residents were less likely to have received booster shots than other ZIP codes with similar education levels and age composition. Strategies used successfully in these communities during the initial vaccine rollout, including outreach efforts and establishment of convenient venues for vaccination, need to be continued during the rollout of boosters to close coverage gaps. MA should also expand efforts to increase vaccine and booster uptake in lower-educated communities regardless of racial composition.

Our study has some limitations. First, our numerator and denominator data come from different sources. People may misreport their ZIP code at the vaccination site or may move to another ZIP code since receiving the vaccine. The population denominator data are estimated for ZIP codes based on aggregation of Census tract-level ACS estimates and under the assumption that the population for 2020 was similar to the population for 2015-2019. Second, sampling error in the denominators results in uncertainty in coverage estimates for specific ZIP codes. Third, a small percentage of MA residents did not provide ZIP codes when they received their primary series or booster vaccine. Fourth, we excluded 167 ZIP codes because they were non-residential or institutional ZIP codes, including PO Boxes, universities, and businesses with dedicated ZIP codes. Exclusion of these ZIP codes yielded a data set with greater comparability across units; however, some MA residents were not represented in our analysis. Due to these exclusions – particularly of university-specific ZIP codes – our totals for vaccination and booster coverage differ somewhat from published statewide estimates. Fifth, we further excluded the smallest ZIP codes (n=63) representing 1% of the population as the estimates were unstable due to very small denominators. Sixth, we lacked individual-level data that would enable inferences on the experiences of essential workers, individuals with different income and education levels, and of different race/ethnicities. Our inferences are therefore restricted to population level statements about people in ZIP codes with different characteristics. Seventh, we do not distinguish between essential workers who were in health care and typically required to be vaccinated and in other sectors. Eight, in assessing ZIP code racial/ethnic composition we used a single metric for percent Black, Latino, or Indigenous, as these populations are concentrated in a relatively small number of MA ZIP codes. Our analysis misses important differences in the experiences of Black, Latino, and Indigenous MA residents.

These limitations should be considered alongside the study’s strengths, namely: the use of official, state-reported data on primary series vaccine and booster shots that was aggregated from data on place of residence collected at vaccination facilities; *de novo* construction of population denominators at the postal ZIP-code level enabling analysis of small-area variation in vaccine coverage not previously reported statewide; assessment of inequities across a range of ZIP code characteristics relevant to the epidemiology of COVID-19 and vaccine uptake; and assessment of inequities stratified by age. Finally, our analysis includes all persons vaccinated through February 2022, including the MA Delta and Winter Omicron waves.

Closing vaccine coverage gaps in MA and in other states will require ongoing concerted effort. The initial MA vaccination campaign in Spring of 2021 was accompanied by daily media coverage and an explicit focus on equity, in part because of a phased roll-out to people with advanced age, co-morbidities, residence in institutional settings, and high-risk occupations, along with programs including the Vaccine Equity Initiative. The late 2021 Delta and Omicron waves refocused public attention on the importance of completing the primary vaccine series and getting a booster shot. By Spring 2022, however, vaccine and booster uptake slowed, as the Winter Omicron wave ended and policymakers encouraged a “return to normal”.

Despite a lull in COVID-19 infection rates, MA is in a precarious position. Large inequities in vaccine coverage among younger adults and children and in booster coverage among all age groups could result in largescale morbidity, mortality, and economic dislocation among vulnerable populations during a future COVID-19 wave. While people who are vaccinated and boosted will likely be protected against severe outcomes, many MA residents have not yet had a booster shot. So long as large gaps in vaccine coverage persist, public health measures such as mask mandates, closures, and capacity limits may have to be reinstated to save lives during a future surge. Our analyses indicate strong geographic and sociodemographic patterns in vaccine and booster coverage, which should allow for targeted outreach efforts that leverage local infrastructure, including school-based immunization, workplace vaccination drives, community-based campaigns, and routine clinical care. Ensuring access and communicating the ongoing importance of vaccination through trusted community sources will be essential. Normalizing vaccination – including the booster shot – must be part of MA’s “return to normal”.

## Data Availability

All data are available online at:
American Community Survey data:
https://www.census.gov/data/developers/data-sets/acs-5year.2019.html
MA vaccination data:
https://www.mass.gov/info-details/massachusetts-covid-19-vaccination-data-and-updates
Replication code is available from the authors upon request.

https://www.mass.gov/doc/weekly-covid-19-municipality-vaccination-report-march-3-2022/download

**Appendix Figure 1.**
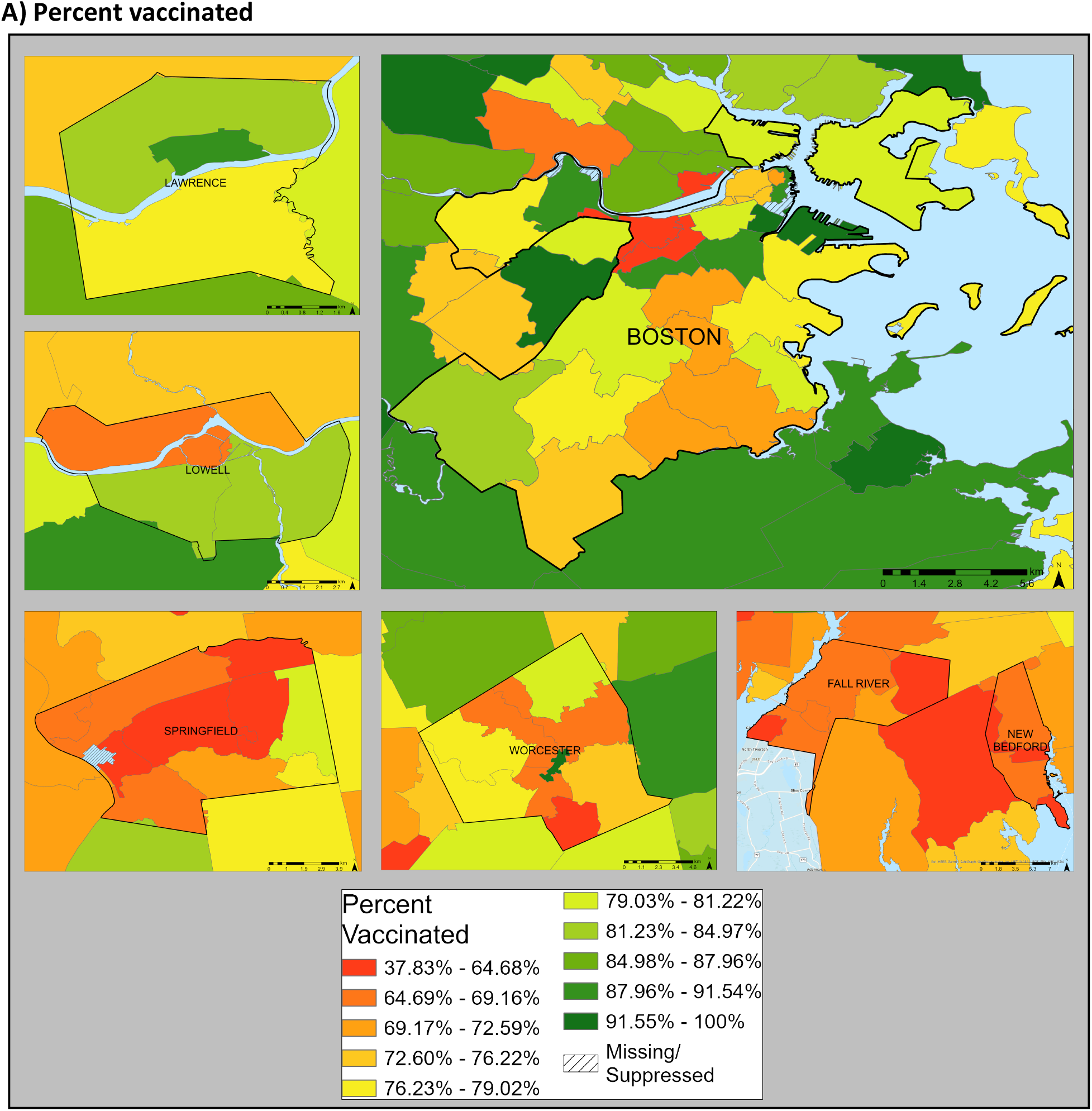
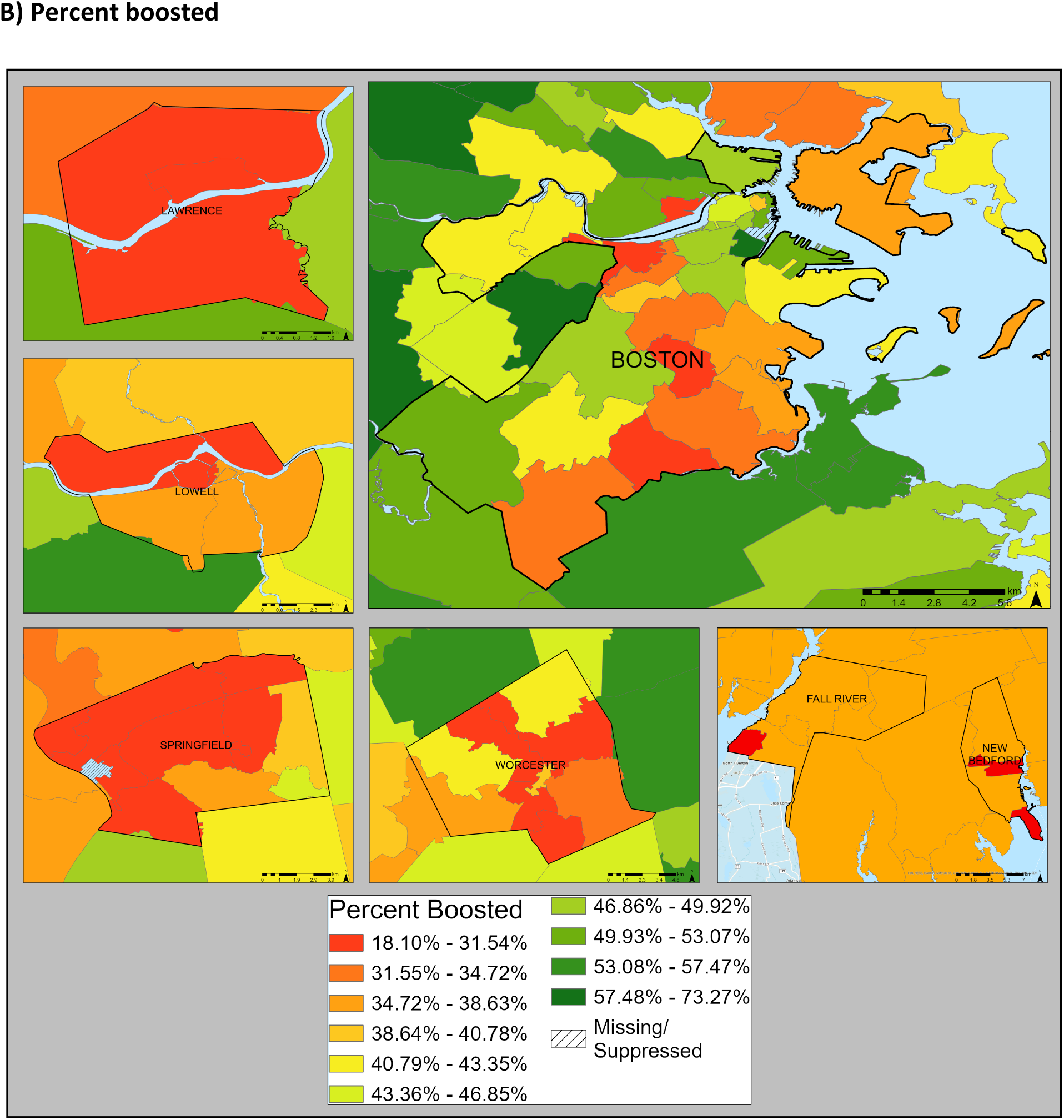
Percent (A) vaccinated and (B) boosted in urban areas.

